# Shared inheritance reveals landscape of somatic and germline cancer risk in *TP53*

**DOI:** 10.64898/2026.04.10.26350605

**Authors:** Hamish A J MacGregor, Jamie R Blundell, Douglas F Easton

## Abstract

Pathogenic variants in *TP53*, the key tumour-suppressor gene underlying Li-Fraumeni syndrome (LFS), are among the best-established causes of inherited cancer predisposition. However, large-scale sequencing has revealed that many apparently pathogenic *TP53* variants detected in blood are the result of somatic clonal expansions, complicating risk interpretation. Using blood-derived whole-exome data from 469,391 UK Biobank participants, we combined variant allele fraction (VAF) with haplotype-sharing analysis to distinguish germline and somatic *TP53* variants. Germline variants were concentrated at sites linked to partial loss of p53 function and lower disease penetrance, whereas classic LFS alleles appeared almost entirely somatic. High-VAF carriers of classic LFS alleles conferred markedly increased risk of haematological malignancy but not solid tumours, consistent with large *TP53*-mutant clonal expansions. The prevalence of somatic clonal expansion also correlated with missense variant pathogenicity, suggesting that somatic activity provides an informative *in vivo* proxy for functional impact. These results provide new insights into *TP53*-associated cancer risk at the population level, demonstrate that somatic rather than germline risk predominates in middle-aged healthy adults and provide a scalable framework for variant classification in large-scale population genomics.

## Introduction

The link between pathogenic mutations in the key tumour suppressor gene *TP53* and Li-Fraumeni syndrome (LFS) – a rare, severe inherited cancer predisposition characterised by multiple early-onset malignancies including bone and softtissue sarcomas and breast cancer ^1^ – is among the oldest and best-established associations in cancer genetics. However, recent advances in sequencing and the rise of population-scale genetic testing have revealed a more nuanced picture of cancer risk among *TP53* carriers.

Genetic screening studies over the last decade have reported that a substantial proportion of people carrying apparently-pathogenic *TP53* mutations, thought to be very highly penetrant, did not present symptoms or a family history suggestive of LFS ^2–8^. Many of these variants were subsequently identified to be the result of *TP53*-driven clonal haematopoiesis (CH), where somatically-acquired mutations in haematopoietic stem cells drive clonal expansions that are detectable in the peripheral blood samples taken for genetic testing. As with other CH drivers, the prevalence of *TP53*-driven CH increases with age ^9^. While CH is benign in most individuals, it is a significant risk factor for myeloid ma-lignancies ^10–14^, and *TP53*-mutant acute myeloid leukaemia (AML) is a particularly aggressive form of the disease with very poor survival ^15,16^. Nonetheless, the risk associated with *TP53*-driven CH is much lower than typical estimates for germline *TP53* variants responsible for LFS (>70% for males and ∼100% in females ^17,18^). Thus, whether a variant is germline or somatic has significant implications for the counselling of individuals and their relatives.

Understanding the distribution of somatic and germline variation across *TP53* may also contribute towards a more refined view of the risks posed by individual *TP53* variants. *TP53* is unusual in that majority of pathogenic variants are missense variants, and the functional interpretation of these variants is not always clear ^19^. Several examples of reportedly ‘lower-penetrance’ variants have been identified, which confer some cancer risk when present in the germline, but do not result in classic LFS phenotypes ^20–23^. However, most studies of germline *TP53* variants rely on small numbers of multiple-case families, making accurate variant-specific risk estimation difficult due to ascertainment bias. For many other *TP53* variants, a lack of empirical data means that their clinical significance remains uncertain. Clonal expansion driven by cancer-related variants is observed across a range of healthy tissues including skin ^24^, oesophagus ^25,26^, lung ^27,28^, and blood ^10,11,29^, and it has been suggested that somatic variation in *TP53* could be used to identify rare, potentially pathogenic mutations ^30^.

Reliably distinguishing between germline and somatically acquired *TP53* variation is challenging, especially in large cohorts with limited sequencing depth and no access to matched tissue samples or longitudinal samples. Most studies primarily rely on variant allele fraction (VAF), since the VAF of somatic variants is usually lower than for germline variants. This can lead to the misclassification of high-VAF (and thus potentially high-risk) somatic variants as germline. In this study, we exploit the scale of the UK Biobank (UKB) to infer germline status from the co-inheritance of genomic segments among carriers, which, alongside VAF data, enables robust classification of 87% of *TP53* pathogenic variants. This approach allows us to quantify the relative prevalence of somatic and germline variation, characterise the distribution of somatic and germline variation across the gene, and to the estimate *TP53*-associated risk in the general population.

## Results

### Shared inheritance and VAF can distinguish somatic and germline *TP53* variation

We identified rare coding variants in *TP53* in 469,391 individuals aged 40-70 in the UKB whole exome sequencing data (WES) using the variant caller Mutect2, with a minimum detectable variant allele fraction (VAF) of 2.7% (Methods). The nonsynonymous variants (n = 1,753) were classified according to a multi-factorial model developed by Fortuno et al. ^31^, which combines computational prediction, *in vitro* functional assays and clinical data to estimate the pathogenicity of *TP53* missense variants (Methods). Carriers predicted to be ‘pathogenic’ or ‘likely pathogenic’ (henceforth ‘PVs’) displayed a strikingly different VAF profile compared with synonymous or predicted-benign carriers (Fig. 1a). The low-VAF range, where variants are more likely to be somatically acquired, is highly enriched for pathogenic variants. Conversely, the germline VAF range (around 50%) is dominated by variants predicted to be benign or synonymous. The prevalence of pathogenic variants clearly increased with age, while the prevalence of benign and synonymous variants remained consistent over time (Fig. 1b).

**Fig. 1.**
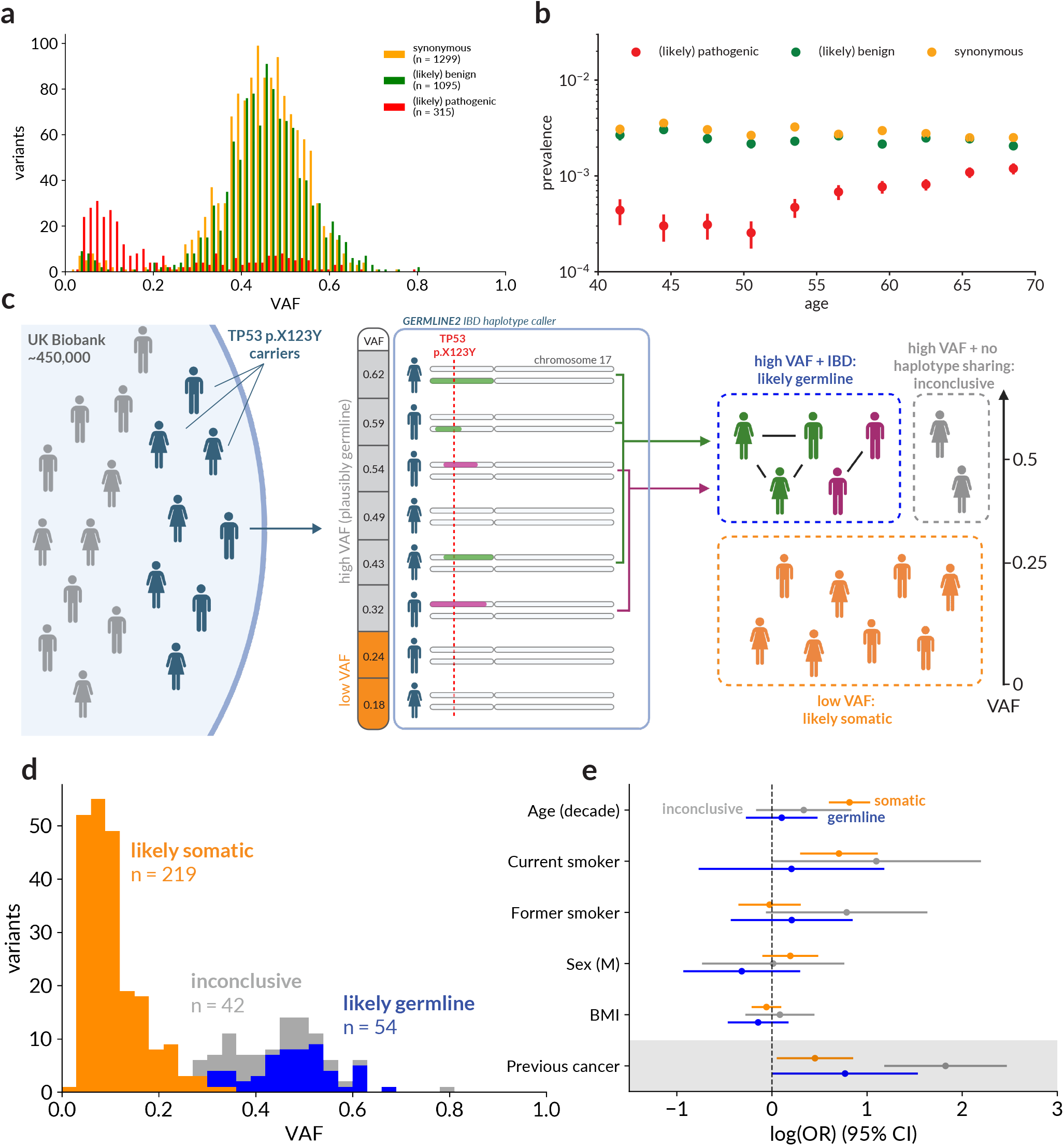
Distinguishing somatic and germline *TP53* variation in UK Biobank. **(a)** VAF distribution of *TP53* coding variants in UKB, split by predicted pathogenicity (pathogenic or likely pathogenic (red), benign or likely benign (green), synonymous (yellow), predictions taken from Fortuno et al. (2021) ^31^**(b)** Prevalence of *TP53* coding variants in UKB across the UKB age range, split by predicted pathogenicity as in (a). **(c)** Schematic showing how *TP53* variants are classified using shared haplotypes. Carriers of a specific *TP53* variant (represented by ‘p.X123Y’) are identified in UKB (left). Phased haplotypes for these individuals are compared pairwise using GERMLINE2 to find shared regions identical by descent (IBD) (middle). For variants where the VAF is high enough that a germline origin is plausible, carriers that share IBD haplotype segments are assumed to be germline, while high-VAF carriers without haplotype sharing are deemed inconclusive. Low-VAF variants are assumed to be somatic (right). **(d)** VAF distribution of pathogenic *TP53* variants, coloured by classification: likely somatic, as determined by the binomial VAF analysis (orange), likely germline, as determined by haplotype analysis (blue) and inconclusive (grey). **(e)** Association between age, smoking, sex, BMI and previous cancer diagnosis with presence of a *TP53* PV, for each classification (somatic, germline, inconclusive). Individuals with a cancer diagnosis before UKB assessment were excluded from the analysis of age, sex, smoking and BMI (unshaded rows). Logarithms are natural (base *e*). Supplementary Table 3 contains full regression results.

To distinguish somatic and germline variants, we combined information from VAF and co-inherited genomic segments. For 70% of PVs (219/315), the VAF indicated a less than 1% chance of germline origin based on a binomial test (Methods), suggesting that the variants were likely to be somatically acquired. For higher-VAF carriers, where a germline origin remained plausible, we applied a haplotype-based approach to detect variants shared identical by descent (IBD). In a relatively homogeneous cohort such as UKB, individuals carrying a rare germline variant are likely to have inherited it from a recent common ancestor, and hence to share extended haplotype segments surrounding the variant. Detecting such IBD sharing among carriers therefore provides strong evidence of germline origin.

Using statistically phased haplotypes from UKB ^32^, we applied the IBD detection algorithm GERMLINE2^33^ to identify groups of *TP53* PV carriers who shared extended haplotype segments overlapping the variant (Fig. 1c, Supplementary Fig. 4, Methods). 54 higher-VAF carriers (17% of total PVs) were found to share an extended haplotype segment with at least one other carrier of the same variant, strongly suggesting that these variants were of germline origin. This left 42 higher-VAF PVs (13%) that could not be conclusively classified (Fig. 1d), either because they did not share identical haplotype segments with other carriers, or because only one carrier was present in the cohort. These ‘inconclusive’ variants may represent either genuine germline alleles without detectable shared ancestry, including *de novo* events, or high-VAF somatic mutations that have expanded to large clonal fractions in blood. Post-zygotic mosaicism could not be identified using this method, but is expected to be exceptionally rare in this cohort (Methods).

### Predictors of pathogenic *TP53* variation

We next investigated the relationship between *TP53* PV prevalence and factors known to influence the development of CH, such as age, smoking and history of cancer. For each of the three variant classes defined above (likely germline, likely somatic and inconclusive), a multivariable logistic regression model was fitted with covariates age, sex, BMI, smoking status, genetic ancestry and whether an individual had been diagnosed with cancer prior to UKB assessment. All three classes of *TP53* PVs were associated with a previous cancer diagnosis, with variants classified as ‘inconclusive’ showing the strongest effect (likely somatic: OR 1.6 (1.1–2.3), *p* = 0.03, likely germline: OR 2.2 (1.0–4.6), *p* = 0.049, inconclusive: OR 6.2 (3.3–11.8), *p* = 3 ×10^*−*8^, Fig. 1e). These associations with previous cancer diagnosis are unsurprising, as germline *TP53* PVs are known to confer substantial cancer risk, and some cancer treatments promote expansion of somatic *TP53*-mutant clones ^34^. However, since the causal relationship between *TP53* variation, smoking and cancer is complex and retrospective analyses are subject to significant selection biases, this could lead to poor estimation of individual effect sizes for other predictors. To isolate the remaining factors from confounding by the effect of cancer treatment, the remainder of the analysis was conducted excluding individuals with a previous cancer diagnosis.

Older age was strongly associated with increased prevalence of ‘likely somatic’ variants (OR 2.3 (1.8–2.8) per decade of age, *p <* 1× 10^*−*10^), further confirming that these variants are likely to be the result of clonal expansion (Fig. 1e). Likely germline variants were not associated with age (OR 1.1 (0.8–1.6) per decade, *p* = 0.59). The odds of carrying a likely somatic or ‘inconclusive’ *TP53* PV were significantly increased in current smokers (somatic OR 2.0 (1.3–3.0) vs never-smokers, *p* = 7 ×10^*−*4^, inconclusive OR 3.0 (1.0–9.0), *p* = 0.047), but not in former smokers. No significant association with smoking was seen for likely germline variants, as expected. No significant association with sex, BMI or genetic ancestry was found for any class of *TP53* PV, and no significant associations were found between any of the covariates and benign or synonymous *TP53* variants (Supplementary Fig. 6).

### Germline variants in UKB show evidence of low penetrance

Among the 55 unique pathogenic *TP53* variants detected in UKB, variants were classified as germline for only six – G262S, R196Q, R181H, G334R, R267Q, and R158H (Fig. 2a). Most fall at positions previously linked to partial loss of p53 function rather than complete inactivation. Previous studies have suggested two of these variants may be associated with reduced disease penetrance (compared with canonical LFS variants): R181H has been reported to retain partial p53 activity and is linked primarily to later-onset breast cancer rather than paediatric LFS-type tumours ^23,35^, while G334R, a founder allele in the Ashkenazi Jewish population, has also been suggested to be associated with lower cancer risks ^21^. For a further three, G262S, R196Q and R267Q, the primary contribution to the pathogenic classification in the Fortuno model is from computational predictions, whereas *in vitro* functional assays assigned a low probability of pathogenicity ^36^, suggesting partial preservation of p53 function.

**Fig. 2.**
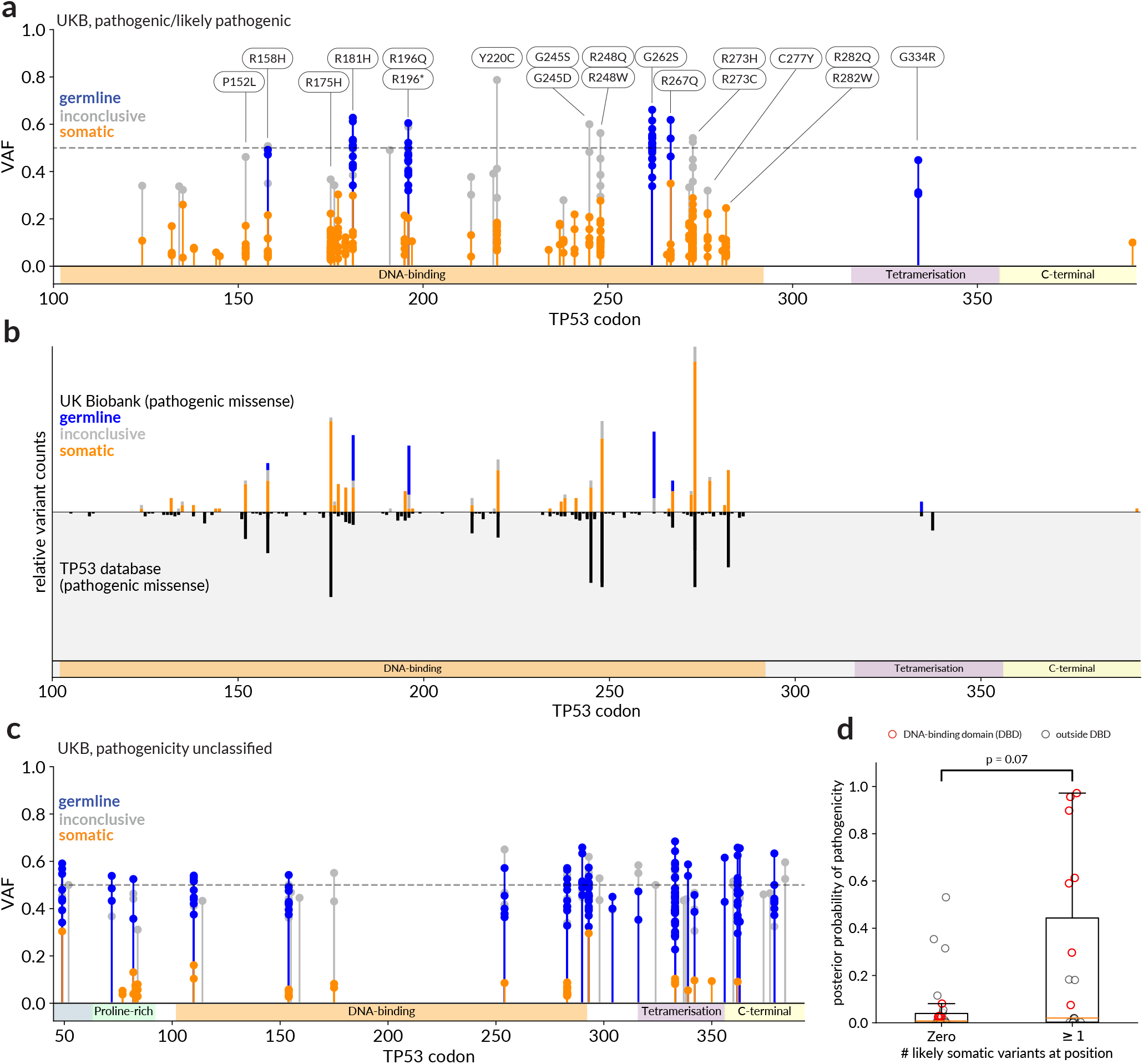
Distribution and cancer risk of *TP53* variants in UK Biobank. **(a)** Position and VAF of all pathogenic or likely-pathogenic *TP53* variants identified in UKB participants. Colours represent classification as somatic (orange), germline (blue) or inconclusive (grey). A selection of variants are labelled with their amino acid substitution, and colours along the x-axis show the domains of *TP53*. **(b)** Relative variant counts for pathogenic *TP53* missense variants in UKB (above, coloured as in (a)) and germline pathogenic missense variants in the IARC *TP53* database (below, black). Variant counts in each case normalised by the total number of eligible variants in the respective database. **(c)** Position and VAF of UKB variants in *TP53* for which no pathogenicity classification was assigned by the Fortuno model. Colours as in (a). **(d)** Box plot comparing Fortuno posterior probabilities of pathogenicity for unclassified *TP53* variants with no likely somatic carriers in UKB against variants with at least one likely somatic carrier. Circles show pathogenicity scores for individual variants, with variants in the DNA-binding domain highlighted in red. P-value derives from a two-sided Mann-Whitney U test.

One variant did not fit this overall pattern. Two individuals carried apparently germline R158H variants, an allele with a well-established association with LFS ^37^. Neither carrier reported a clinical or family history suggestive of LFS. Analysis of background haplotype sharing suggested that the probability of this shared segment arising by chance was less than 5 ×10^*−*4^, given the total number of R158H carriers (n = 11) and the shared segment length of 3.5 cM (Methods).

### Somatic clonal expansion correlated with variant pathogenicity

To test the hypothesis that somatic expansion in *TP53* correlates with germline pathogenicity, we compared the distributions of *TP53* variants found in UKB against the distribution of germline variants in the *TP53* Database (*TP53*db), a public online resource containing more than 50,000 *TP53* variants reported in the literature ^38,39^. Among variants predicted to be pathogenic, there was a striking similarity between the frequency distribution of likely somatic variants in UKB and germline *TP53* variants reported in *TP53*db (Fig. 2b). The top three most frequently observed somatic PVs in UKB (R273H, R175H and R248Q) are also the three most common germline PVs in *TP53*db, and are all well-established canonical LFS alleles. Conversely, at the three positions with the most frequent germline variants (G262S, R196Q and G334R) no low-VAF variants were detected in UKB.

The link between somatic variation and pathogenicity may help us identify further lower-penetrance *TP53* alleles. 45 variants identified in UKB were not given a formal classification by the Fortuno model due to a lack of clinical data ^31^ (Fig. 2c, Supplementary Table 2). We divided these into variants with and without likely somatic carriers in UKB, as determined by the binomial VAF test, and compared their posterior pathogenicity scores from the Fortuno model (based on in vitro and computational predictions only). Across the full *TP53* coding sequence, variants with likely somatic carriers tended to have higher pathogenicity scores than those without evidence of somatic expansion, although this difference did not reach statistical significance (Mann-Whitney U-test, two-sided, *p* = 0.07; Fig. 2d). Inspection of the data suggests that this signal is primarily driven by variants within the DNA-binding domain (codons 102–292), which is known to harbour the majority of pathogenic *TP53* variants. No carriers were observed for the tetramerisation-domain variant R337H, a founder allele prevalent in Brazil and associated with reduced-penetrance LFS-like phenotype ^20^. Given that R337H arises from a C>T transition, for which the underlying somatic mutation rate is typically high ^40^, somatic occurrences might be expected at this position. Most variants with at least somatic carrier were also found in the germline in UKB, suggesting that, if pathogenic, these are lower-penetrance variants similar to the variants discussed above.

### Haematological cancer dominates risk for *TP53* carriers in UKB

We assessed the effect of *TP53* pathogenic variant (PV) carrier status on incident cancer risk using Cox regression, with adjustment for genetic ancestry, sex, body mass index, and smoking status (Fig. 3). Hazard ratios (HRs) were estimated separately for each variant class (likely somatic, inconclusive, and likely germline). A significant increase in all-cancer risk was observed for likely somatic and germline variants (somatic HR 1.5 (1.1–2.1), *p* = 0.017, germline HR 2.1 (1.1–3.9), *p* = 0.02), and an increase was observed for inconclusive variants that did not reach significance (inconclusive HR 2.0 (0.9–4.5), *p* = 0.08).

**Fig. 3.**
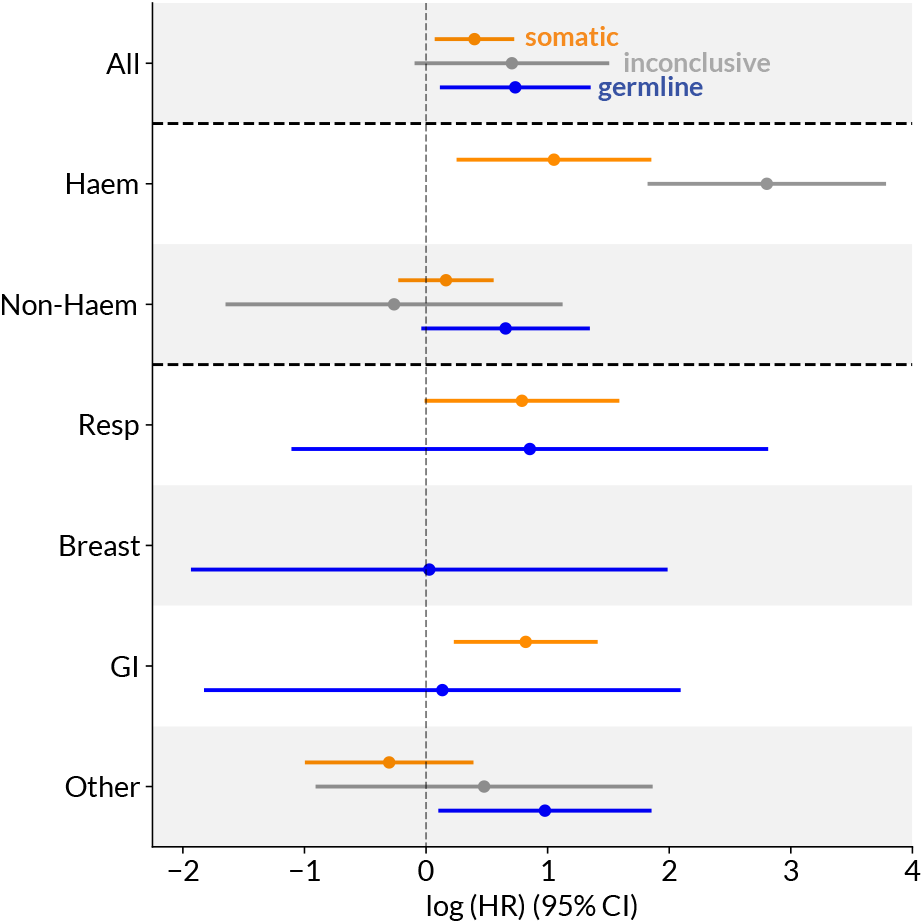
Incident cancer risk in UKB *TP53* carriers. Estimates of loghazard ratio (natural log, with 95% CI) for risk of incident cancer associated with carrying a *TP53* variant classified as likely somatic (orange), inconclusive (grey) or likely germline (blue), derived from a Cox proportional-hazards model. Top row shows all-cancer risk, subsequent rows segregated by cancer site: haematological (Haem), all non-haematological (Non-Haem), respiratory (Resp), breast, gastrointestinal (GI) and any other site. Missing rows indicate that no case-carriers exist in UKB for that combination of cancer site and variant class. Full results in Supplementary Table 4.

The pattern of risk, however, differed sharply by variant origin. For both somatic and inconclusive carriers, the signal was driven almost entirely by haematological cancers, with effect among inconclusive carriers particularly striking (somatic HR 2.9 (1.3–6.4), *p* = 0.010, inconclusive HR 16.5 (6.2–44.0), *p* = 2× 10^*−*8^). The high risk of haematological malignancy, but not of solid tumours, is markedly different from the pattern seen in LFS resulting from germline *TP53* variants. These high-VAF ‘inconclusive’ variants are therefore likely to represent large, somatically acquired *TP53*-mutant clones whose expansion drives a substantial rise in blood cancer risk.

For germline variants, conversely, no prospective cases of haematological cancer were observed in carriers. A weak association was seen with cancer aggregated across sites outside the four main categories (i.e. not haematological, respiratory, breast or gastrointestinal), including cases of prostate and brain cancer (HR 2.7 (1.1–6.3), *p* = 0.03). However, the case counts for all sites were small and confidence intervals wide. One additional significant association was observed between likely-somatic carriers and gastrointestinal cancer (HR 2.3; (1.3–4.1), *p* = 0.007), while the association with respiratory cancer was marginally non-significant (HR 2.2; (1.0–4.9), *p* = 0.05) – these findings are discussed further below.

## Discussion

These findings provide a refined view of *TP53*-associated cancer risk in the general population, using data from a large population-based cohort. Clinical screening has traditionally focused on identifying germline pathogenic variants responsible for LFS-like susceptibility, whereas somatic mutations detected in blood have generally been regarded as comparatively benign. In the UK Biobank cohort, we found that true germline LFS alleles are, as expected, exceptionally rare, reflecting the strong negative selection against high-penetrance *TP53* mutations. However, somatic variants occurring at these same canonical sites were relatively common and conferred a significant cancer risk. This excess risk was driven almost entirely by haematological malignancies, for which *TP53*-mutant disease is characterised by treatment resistance and extremely poor prognosis ^16,41^. The strongest association was seen for variants with the highest VAF, consistent with our current understanding of the progression from CH to malignancy ^42^. This suggests that a purely VAF-based approach to distinguishing somatic and germline variants, where higher-VAF somatic variants may be misclassified as germline, will likely underestimate the haematological cancer risk associated with *TP53*-driven CH.

Evidence for an association between CH and non-haematological cancer remains inconsistent. While we observed an association between *TP53*-driven CH and gastrointestinal cancer, previous studies have reported conflicting results – some analyses in UKB have found elevated risk for gastric and colorectal cancer among CH carriers (any driver) ^43–45^, whereas comparable analyses in the TOPMed dataset found no association ^46^. Residual confounding by smoking or other CH-promoting environmental exposures may contribute to these findings.

The variants identified as germline appeared to be over-represented by variants with prior evidence of being ‘lower penetrance’. This is not surprising since the cohort recruited middle aged adults, leading to strong selection against higher risk variants. The risk estimates cannot be compared with previous estimates for LFS (which are based on all ages and potentially subject to ascertainment biases). The comparatively lower disease risk associated with germline variants found here should not be interpreted as implying that ‘lower-penetrance’ germline alleles are benign: although the germline risks estimated here appear lower than for individuals with LFS, the point estimate of HR observed here would still correspond to a substantially elevated cumulative cancer risk between ages 50-80 (males 59% vs 37% baseline, females 48% vs 30%, using CRUK incidence statistics ^47^).

Questions remain regarding which specific non-LFS missense variants confer risk, as the present study lacked power to analyse individual *TP53* variants. As others have suggested ^30^, we found that the presence of expanding somatic clones correlates strongly with likely pathogenicity of variants, suggesting that somatic selection may serve as an in vivo indicator of functional impact. This relationship may be weaker for variants outside the DNA-binding domain. Future deep sequencing studies, with greater power to resolve low-VAF clones, could further characterise this spectrum and may be able to identify further rare pathogenic alleles. Quantitative aspects of this relationship are also still unclear, for instance, whether the correlation between the prevalence of LFS alleles and CH drivers is primarily due to correlated mutation rates or fitness advantages. It is important to emphasise that cancer risk in PV carriers is likely to be influenced not only by the variant but also by other genetic and lifestyle factors. Further analyses in larger cohorts will be needed to validate these variant-specific associations and, ultimately, provide reliable variant-specific cancer risk predictions.

## Methods

### Variant calling

Variants were called from the UKB exome CRAM files using the somatic variant caller Mutect2 in tumour-only mode ^48^. Additional filters were applied to exclude potential sequencing artefacts: Any variant with a total coverage of fewer than 10 reads was excluded. Variants were only retained if supported by at least 3 variant reads, with at least one on each of the forward and reverse strands (similar to the UKB somatic variant calling strategy used by Tran et al. ^49^). The strand requirement was lifted if the Phred-scaled strand bias score generated by Mutect2 was at least 30 (indicating a less than 1 in 1000 chance of strand bias), in order to avoid excluding variants where the total coverage was significantly higher on one strand than the other. Variants with a total prevalence of more than 0.1% in UKB were excluded. After filtering, 1,753 nonsynonymous coding *TP53* variants were identified. The lowest detectable VAF was 2.7%. The *TP53* transcript NM_000546.6 (ENST00000269305.9) was used for all analysis. The DNA-binding domain was taken to be between codons 102 and 292 inclusive.

### Pathogenicity of *TP53* missense variants

The nonsynonymous variants were classified using a multifactorial model developed by Fortuno et al. (2021) ^31^. 315 (18%) were deemed pathogenic or likely pathogenic, 1,095 (62%) benign or likely benign and 82 (5%) of uncertain significance. In the remaining 261 cases (15%), the model was not applied as the variant had not been observed in at least one of the ‘clinical’ dataset used to determine pathogenicity.

The Fortuno model only assessed the pathogenicity of missense variants, not truncating variants or large insertions or deletions. Three examples of these variants were seen in UKB:

- Stop-gain (nonsense) mutation R196* (n = 2 in UKB). This variant has been observed frequently in both LFS and in tumours, and is well-known to be pathogenic. Assigned pathogenic.
- Frameshift mutation T125Hfs*24 (n = 2). This nonsense variant is considered pathogenic in ClinVar and observed in COSMIC, although with only one observation in each dataset. Assigned pathogenic.
- Long inframe deletion (GGCTGGTGCAGGGGCCGCCGGTG-TAGGAGCT>G) at codon 78-88 (n = 157). Only observed at low VAF and deemed of uncertain significance in ClinVar based on four observations. Assigned ‘variant of uncertain significance’.

The 315 pathogenic or likely pathogenic variant calls are listed in Supplementary Table 1.

### UKB cohort and cancer status

The total number of UKB participants included was 454,758 (excluding individuals with no exome sequencing data available at time of analysis, or unknown age at enrolment). 32,902 individuals had at least one cancer diagnosis before their UKB blood sample was taken (either self-reported or from the linked cancer registry data), including 20 for whom the age at diagnosis was unknown. 47,447 individuals had no history of cancer but were subsequently diagnosed after enrolling in UKB. At the time of analysis, the latest available cancer registry data was from 15 March 2022.

For the site-specific analysis, incident cancers were divided into five main groups: gastrointenstinal (n = 11,857), respiratory (n = 4,322), lymphoma and leukaemia (5,853), breast (including carcinoma in situ, n = 15,667) and any other site (n = 29,962).

Body mass index (BMI, height/weight^2^) was calculated from height and weight measured at assessment. Height or weight information was missing for 1,832 participants. Genetic ancestry was estimated from array genotype data using *iAdmix* ^50^, which calculated the probability of belonging to one of four main ancestry groups (European, South Asian, East Asian, African). The probability threshold for a definitive classification was 80% (70% for South Asian), if this threshold was not met, the individual was classified as Mixed (full details in Ficorella et al. (2025) ^51^).

Smoking was encoded as a three-level ordered categorical variable: ‘never smoked’ (including people who did not smoke currently but had selected ‘just tried once or twice’ for past smoking), ‘former smoker’, and ‘current smoker’.

### Binomial analysis of VAF

Binomial analysis of VAF was used to identify likely somatic variants. Read counts for germline variants would be expected to follow a binomial distribution Bin 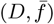, where *D* is the sequencing coverage and mean VAF 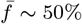. VAF distributions for synonymous and benign variants followed binomial distributions closely (Supplementary Fig. 1)., given a mean VAF estimated from the mean of benign or synonymous variants (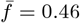 in both cases) and a distribution of sequencing depth *D* matching the observed distribution for *TP53* in UKB. The fact that the observed mean VAF is below 50% is likely the result of slight general alignment bias towards the reference allele, commonly encountered in sequencing studies ^52^. There were a few low-VAF outliers among both benign and synonymous variants that are unlikely to represent true germline variants – variants with VAF < 20% were excluded for the purposes of estimating the mean VAF.

For PV carriers, each variant was assigned a binomial p-value (one -sided) for the null hypothesis that the variant is germline. A significance threshold of *p <* 0.01 was chosen, corresponding to an expected false positive rate of approximately one variant among the 96 examples where null hypothesis was not rejected.

### Phased genotypes

Phased genotypes were obtained from UKB (field 22438). Statistical phasing had been performed by Bycroft et al ^53^ for 487,442 UKB participants using the *SHAPEIT3* algorithm ^54^ with the 1000 Genomes Phase 3 dataset ^55^ as a reference panel. Bycroft et al. estimated that the switch error rate was around 0.23%. *qctool* v2.0.8 was used for processing of phased genotype data.

### Identification of shared haplotype segments

Haplotype segments shared between individuals were identified using the *GERMLINE2* algorithm (Saada et al. (2020) ^33^) paired with a recombination map created by deCODE Genetics ^56^. All *TP53* PVs with at least two UKB carriers were analysed, accounting for 302 of 315 *TP53* PV carriers in UKB. For each eligible variant, *GERMLINE2* identified pairs of haplotypes belonging to carriers of that variant that shared an identical segment (a) at least 1.5cM in length (see next section), and (b) overlapping the *TP53* PV. The extent of the search area was chromosome 17p. After evaluating all shared pairs for a given variant, we constructed an undirected graph with nodes representing individuals (separate nodes for the paternal and maternal chromosome) and edges representing shared haplotype segments. Taking the connected components of this graph (computed with Python *NetworkX* v3.5) yielded groups of individuals sharing a putative IBD segment (Supplementary Fig. 4). Membership of one of these groups therefore indicated a high likelihood of germline origin for the *TP53* variant.

In *n* cases, both of an individual’s chromosomes were found to share haplotypes in separate groups. Since VAF indicated that the variant was heterozygous, it is likely that one of these identical regions represents a false positive. In each case, the individual was assigned to the larger group, and the variant was classified as likely germline. In one Y220C carrier (VAF 79%), GERMLINE2 identified a run of homozygosity overlapping TP53 (not shared with other Y220C carriers); this copy-neutral loss of heterozygosity had been previously identified by Loh et al. ^57^. This behaviour is not indicative of germline origin and this variant was classed as ‘inconclusive’.

### Minimum length of shared haplotype and false positive rate

The choice of minimum shared haplotype length represents a balance between sensitivity and specificity. Shorter thresholds increase the risk of false positives arising from coincidental haplotype sharing, whereas a longer threshold reduces false positives but may miss true, more distant germline relationships. The expected frequency of false positives (i.e. pairs or groups of individuals who share identical haplotype segments by chance overlapping the *TP53* locus) also depends on the local recombination rate, the population structure of the UKB cohort and the size of the groups being compared. To estimate the false positive rate for a given minimum shared length, we drew 400 sets of random UKB individuals (without replacement) from UKB, organised into groups the same size as the number of carriers for each *TP53* PV. For each group of random ‘carriers’, we used GERMLINE2 to identify shared haplotype segments for different minimum shared lengths. The relationship between the mean proportion of individuals carrying a shared segment and minimum segment length is shown in Supplementary Fig. 2a). The minimum length was chosen to be 1.5cM, corresponding to an expected false positive rate of 0.02 per individual, or ∼6 false positives among the UKB *TP53* PV carriers. The per-individual chance of a false positive in a group increases linearly with the number of individuals in the group (since the number of comparisons per individual increases linearly with group size, Supplementary Fig. 2b), the expected number of false positives in a given group therefore increases quadratically with group size. In the UKB *TP53* analysis, six likely false positives were identified (three pairs of individuals who share identical haplotype segments overlapping *TP53*, but where the VAF is too low to indicate germline origin), in agreement with predictions (Supplementary Fig. 3). It is possible, however, that there are also further false positives at higher VAF.

### Post-zygotic mosaicism

It was not possible in this study to identify postzygotic mosaic (PZM) *TP53* variants, in which a mutation arises during embryonic development and is therefore present in some, but not all, tissues. Such mosaic variants are thought to account for a proportion of individuals with Li-Fraumeni-like phenotypes (although published estimates vary widely ^5,6,8,58,59^), and some carriers may remain asymptomatic. Both the VAF and the associated cancer risk can vary considerably depending on the developmental timing of the mutation. However, PZM events are expected to be even rarer than *de novo* germline mutations, and individuals carrying them are unlikely to be well represented in UK Biobank due to survival bias. Together, these factors make it improbable that post-zygotic mosaicism contributes materially to the effects observed here.

### Statistical methods

A standard logistic regression analysis was used to assess factors that may predict *TP53* carrier status, implemented in Python *statsmodels* v0.14.4^60^. Previous cancer diagnosis, smoking status and ancestry group were included as Boolean variables. ‘European’ was the reference group for ancestry, and ‘never smoked’ was the reference group for smoking. Z-scaling was applied to the BMI data (mean = 27.4 kg/m^2^, *σ* = 4.8 kg/m^2^). Age (at assessment centre) was divided by 10 to give an odds ratio per decade. The significance threshold was *p <* 0.05. After excluding individuals with missing smoking, BMI, or ancestry information, the total number of observations was 450,582. After excluding individuals with a history of cancer, the remaining number of observations was 418,011.

Survival analysis was performed using a standard Cox proportional hazards model, with regression implemented in Python using the *lifelines* package (v0.30.0) ^61^. Any individual with a diagnosis of cancer before UKB assessment was excluded (details above). Models were left-truncated (i.e. individuals entered the study at their age at UKB assessment). Individuals who died before a cancer diagnosis were censored at the age of death (available from the UKB death registry linkage), and individuals with no record of death or incident cancer were censored at their age on the 15 March 2022, the latest linkage to the cancer registry. The median follow-up was 12.9 years. Treatment of covariates was identical to the logistic regression analysis described above. Individuals with missing data for any covariate were excluded. The threshold for significance was *p <* 0.05.

### The *TP53* database

Tables of reported germline variants were retrieved from the NCI *TP53* database (R21, Jan 2025, https://tp53.cancer.gov) on the 6th August 2025. This dataset contains information on individuals that are carriers of a *TP53* germline variant and families in which at least one family member has been identified as a carrier of a germline variant in the *TP53* gene. Criteria for inclusion are the following: a) individuals carrying a sequenced *TP53* germline variant, affected or not by a cancer, b) individuals affected by a cancer and belonging to a family in which at least one family member has been identified as a carrier of a germline variant in the *TP53* gene ^38^. For the variant distribution analysis (Fig. 2), each variant was only counted once per family, irrespective of the number of carriers in that family present in the database. 834 distinct families carried a missense germline *TP53* variant.

Although the database includes individuals without cancer, it is strongly enriched for LFS and LFS-like conditions: of the 834 families carrying a missense germline variant (irrespective of predicted pathogenicity), 353 met either classic or Chompret LFS criteria, a further 112 displayed Li-Fraumeni-like symptoms not meeting the strict clinical definition, and a further 143 reported non-LFS-like family history of cancer. Among the carriers without reported family history of cancer, a number displayed clearly LFS-like symptoms (e.g. early breast cancer or soft tissue sarcoma) – such individuals could, for instance, be *de novo TP53* carriers. As such, families were not excluded on the basis of lack of family history of cancer or LFS status. As for UKB, predicted pathogenicity was assigned using the Fortuno classification model already described. Further information on the *TP53* database is found at https://tp53.cancer.gov.

## Data Availability

Data is available online at https://github.com/the-blundell-lab/SomaticGermlineTP53Risk, except data which is restricted from the public by UK Biobank.

## Acknowledgements

We thank Arnold Levine for discussions during the development of ideas for this work. We thank members of the Blundell Lab for input throughout the project. This research has been conducted using the UK Biobank Resource under application number 28126. JRB is funded by a UKRI Future Leaders Fellowship and by the CRUK Cambridge Cancer Centre. DFE is supported by the NHS in the East of England through the Clinical Academic Reserve.

## Author Contributions

The project was jointly conceived by JRB and DFE. Variant calling and analysis was performed by HAJM. The manuscript was written by HAJM and edited by all authors.

## Data Availability

UK Biobank WES and phased haplotype data was obtained and processed on the UKB Research Analysis Platform (RAP). Individual-level UK Biobank data is available to approved researchers, and individuallevel outputs will be returned to UKB upon publication. Tables of variant pathogenicity predictions are available from Fortuno et al. (2021), DOI https://doi.org/10.1002/humu.24264. The deCODE average recombination map was downloaded from the UCSC Genome Browser (https://genome.ucsc.edu/). The *TP53* database is accessible at https://tp53.cancer.gov.

## Code Availability

Code used for this analysis can be found at github.com/the-blundell-lab/SomaticGermlineTP53Risk.

## Supplementary Material

**Supplementary Fig. 1.**
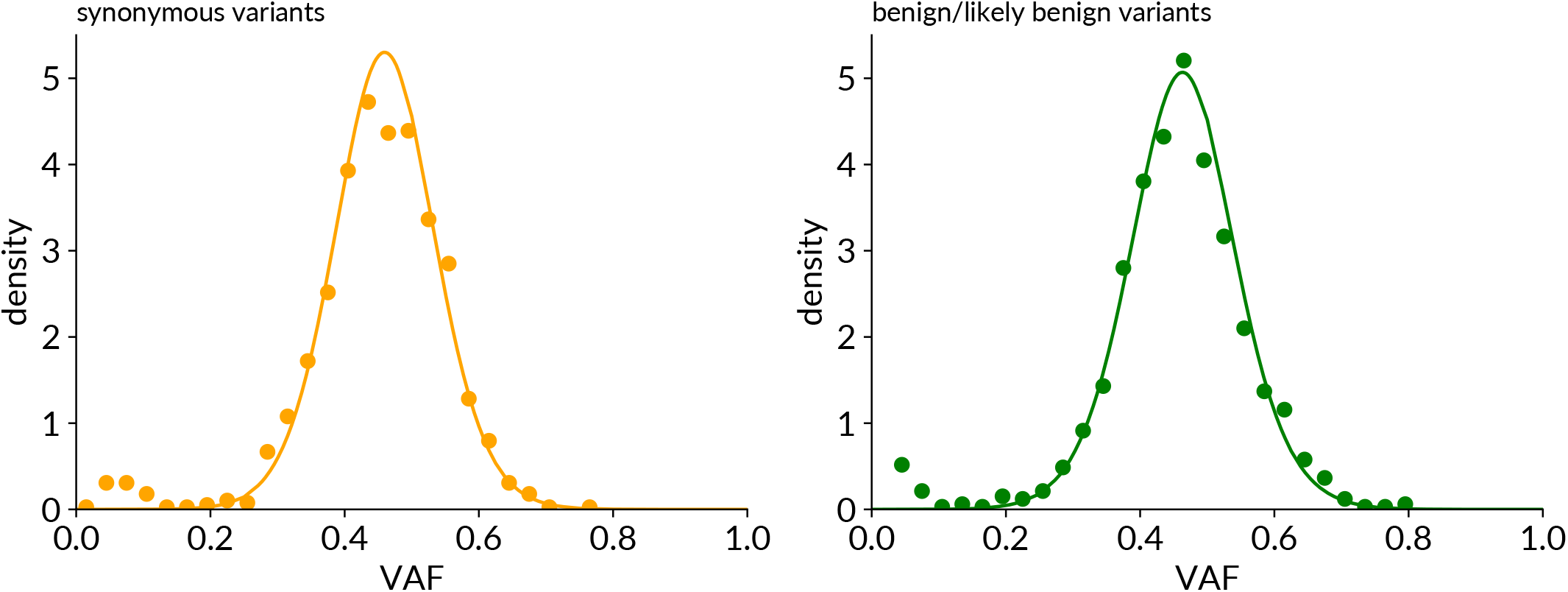
Binomial VAF distribution for germline variants. Points show VAF distributions of synonymous (left) and benign/likely benign missense (right) TP53 variants in UKB, compared with expected distributions derived from a binomial model Bin 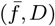 (solid lines). The mean 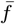 was 0.46, and the binomal was integrated over a distribution of sequencing depth *D* matching the observed sequencing depth in the UKB WES data.

**Supplementary Fig. 2.**
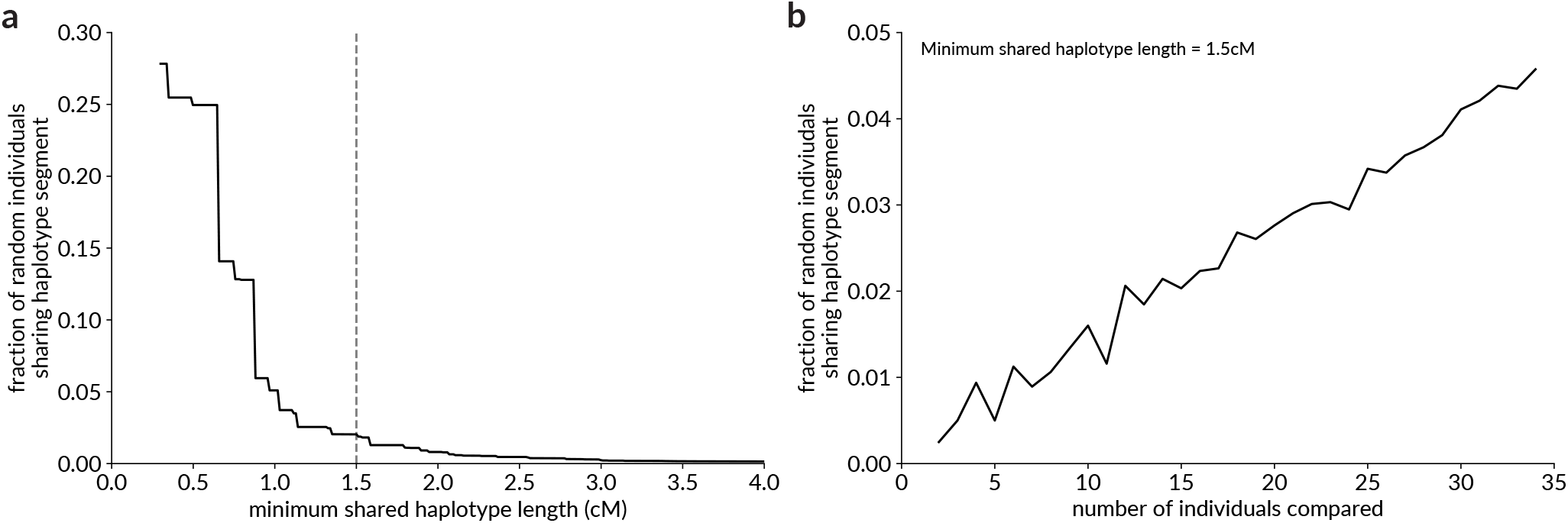
False positive rate for shared haplotype analysis. **(a)** Individuals were drawn at random (without replacement) from UKB and compared in groups equivalently sized to the TP53 PV carriers, repeated 400 times. Solid line shows the mean fraction of random individuals that share a haplotype segment with at least one other individual, for different values of the minimum allowed segment length. Dashed line shows the minimum segment length chosen for the main analysis (1.5cM). **(b)** Fraction of randomly chosen UKB individuals sharing a haplotype segment (>1.5cM) against the size of the comparison group. The increase in the per-person chance of haplotype sharing increases linearly with group size, as expected.

**Supplementary Fig. 3.**
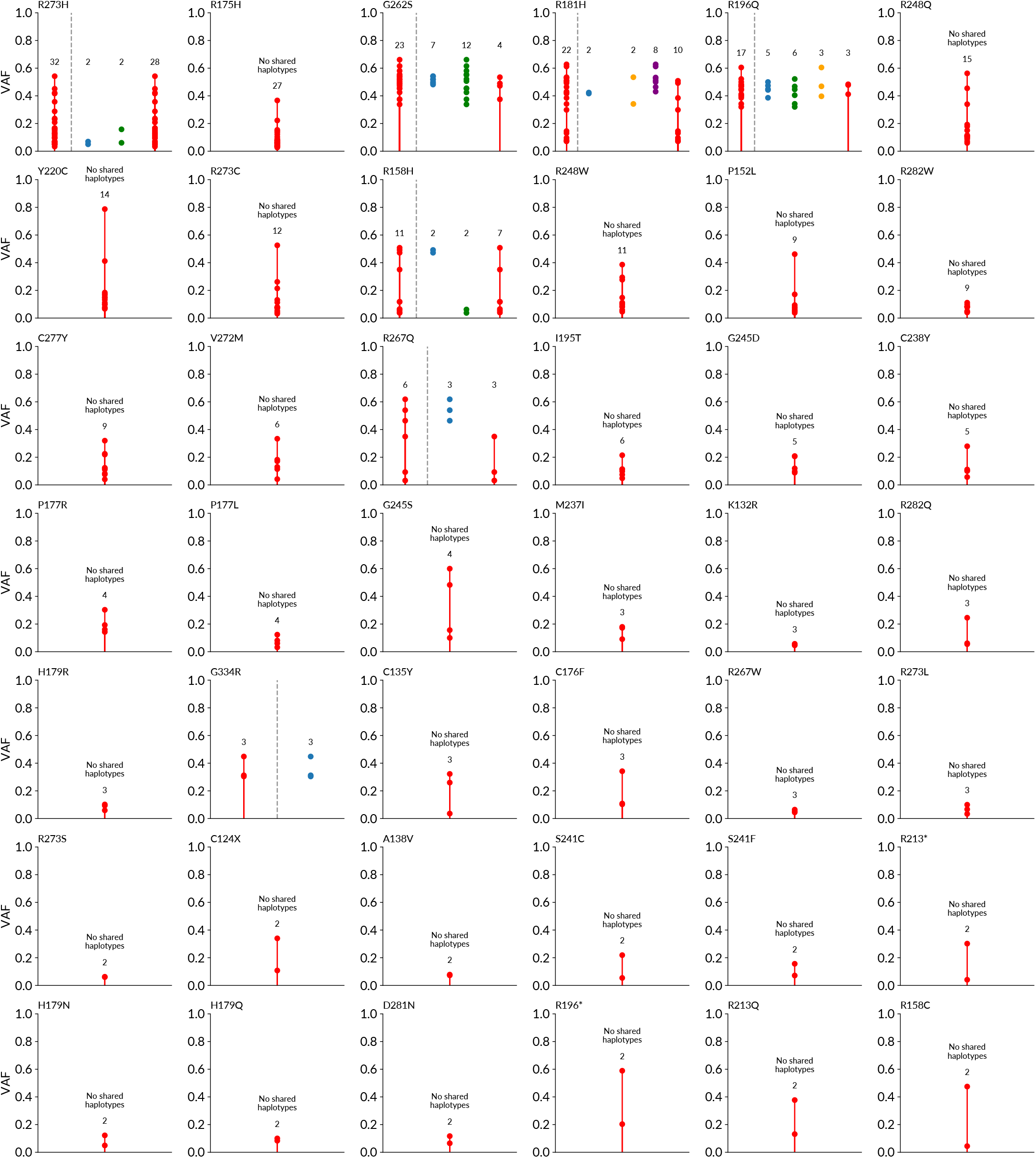
Results of GERMLINE2 haplotype sharing detection. For each TP53 PV with at least two UKB carriers: leftmost column shows VAF of all carriers. If present, columns on the right show all carriers divided into groups sharing IBD haplotype segments, with the number of carriers in each group shown above. Rightmost (red) column, if present, shows remaining carriers not assigned into IBD groups. Likely ‘false positive’ groups, with low VAF, are visible among R273H and R158H carriers.

**Supplementary Fig. 4.**
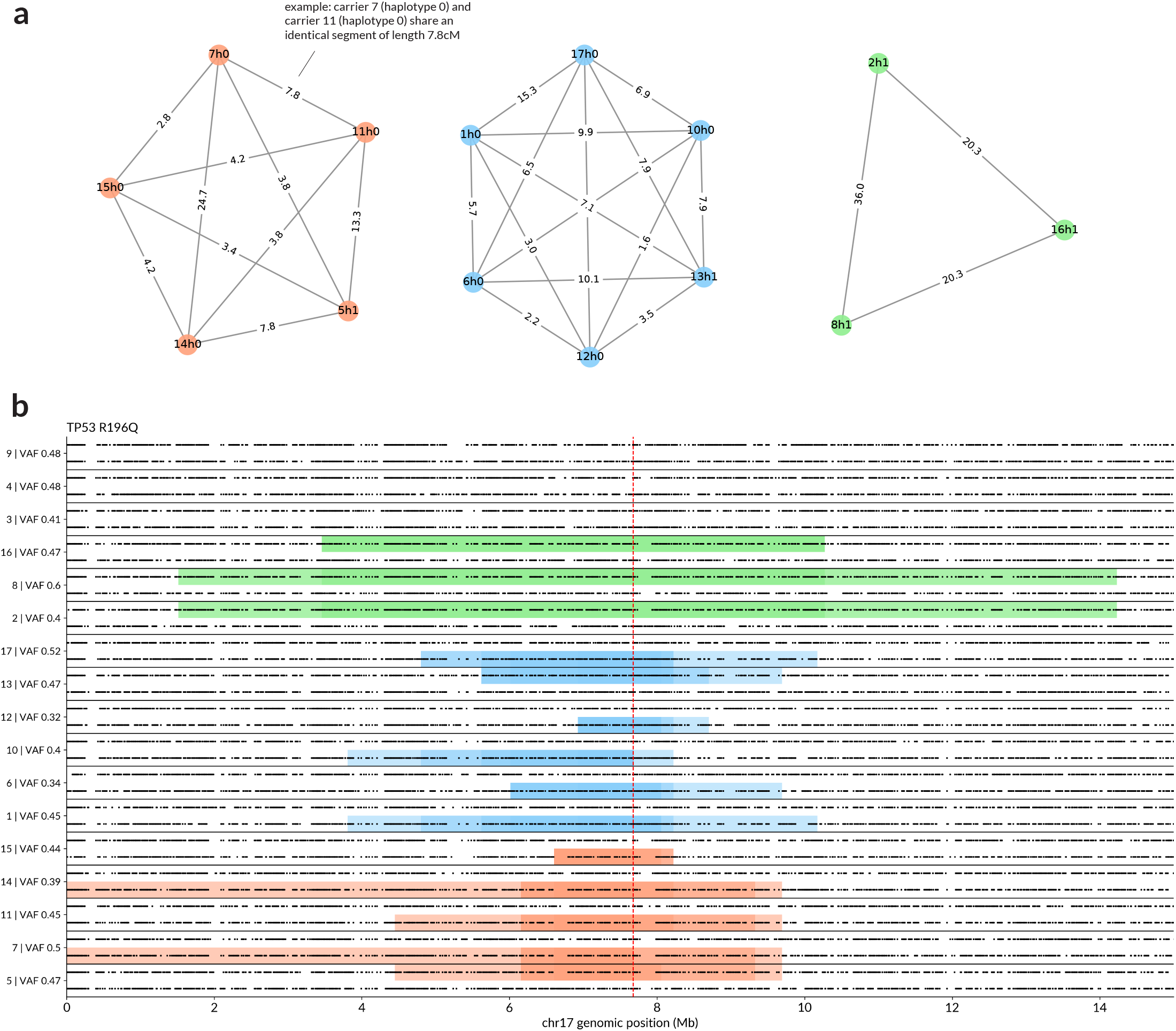
Example of haplotype sharing among 17 TP53 R196Q carriers. **(a)** Undirected graph showing connected components for carriers of the TP53 R196Q variant (n=17 total). Nodes represent individual haplotypes, labelled with the (arbitrary) ID of the carrier and haplotype label (h0 or h1). Edges indicate that two haplotypes share an identical region (>1.5cM) overlapping *TP53* R196Q, and are labelled with the length of this region (in cM). Haplotypes that do not share regions are not shown. **(b)** Phased haplotypes for all 17 *TP53* R196Q carriers in UKB, with points representing heterozygous or homozygous-alternate SNPs on chromosome 17p. Shaded regions indicating identical shared haplotype segments between individuals. Regions shared between more individuals are shaded darker. Colours indicate different connected components of the haplotype sharing graph (i.e. independent lineages), as shown in (a). Red dashed line shows the position of TP53 R196Q. Carrier ID (1-17) and VAF are shown on the y-axis.

**Supplementary Fig. 5.**
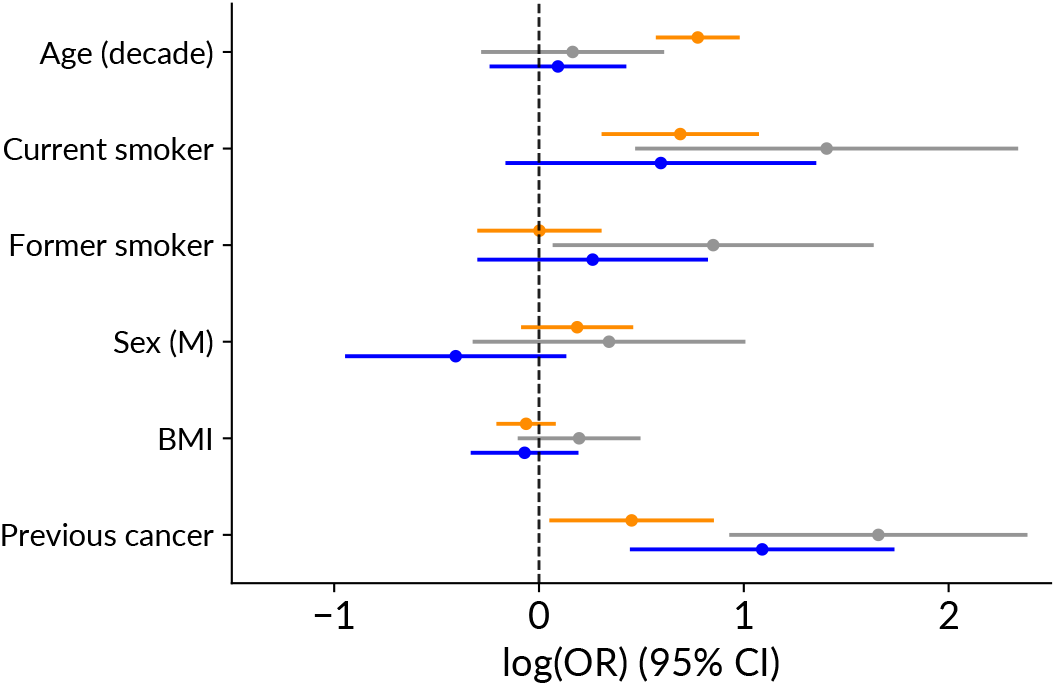
Predictors of pathogenic TP53 variation, including individuals with history of cancer. Association between age, smoking, sex, BMI and previous cancer status and presence of a TP53 variant, for each classification (as in main Fig. 1e). Likely somatic: orange, inconclusive: grey, likely germline: blue. In this analysis, individuals with previous history of cancer are included.

**Supplementary Fig. 6.**
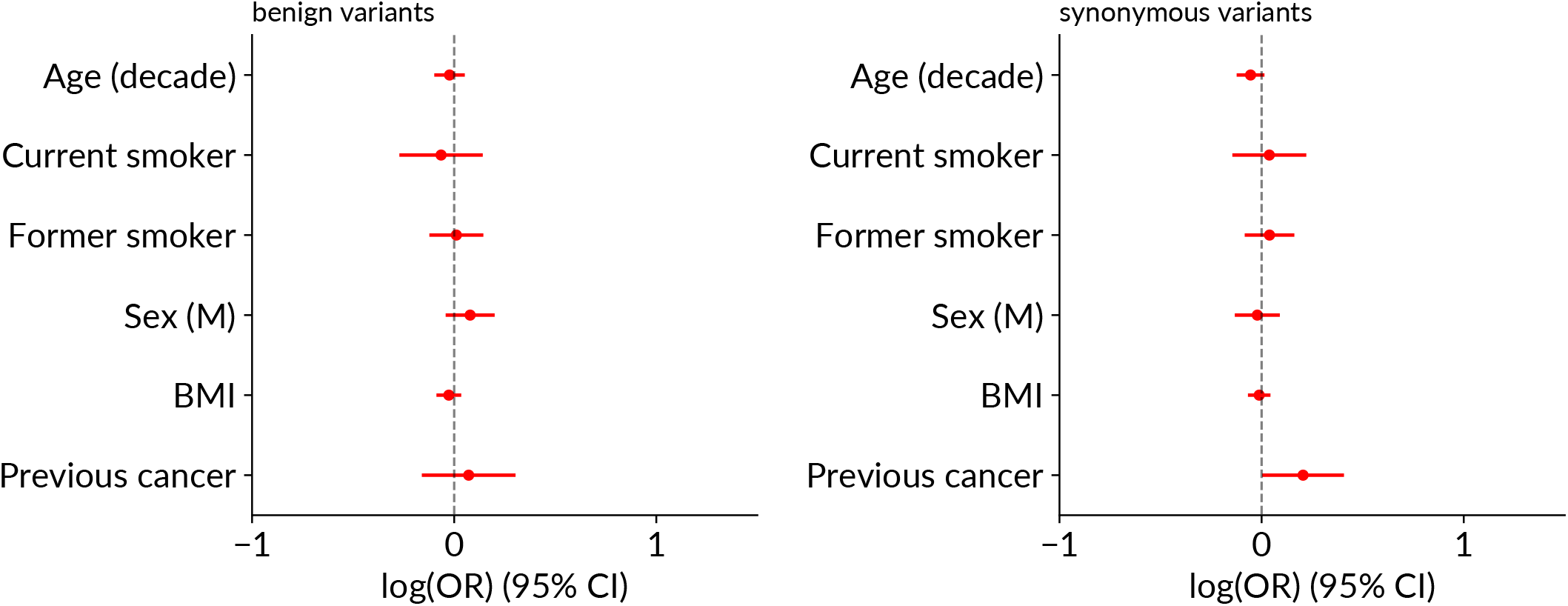
Predictors of benign and synonymous TP53 variation. Association between age, smoking, sex, BMI and previous cancer status and presence of a TP53 variant, for benign variants (left) and synonymous variants (right).

## References

1. F. P. Li and J. F. Fraumeni. Soft-tissue sarcomas, breast cancer, and other neoplasms. A familial syndrome? Annals of internal medicine, 71(4):747–752, 1969. ISSN 00034819. doi: 10.7326/0003-4819-71-4-747.

2. Bradford Coffee, Hannah C. Cox, John Kidd, et al. Detection of somatic variants in peripheral blood lymphocytes using a next generation sequencing multigene pan cancer panel. Cancer Genetics, 211, 2017. ISSN 22107770. doi: 10.1016/j.cancergen.2017.01.002.

3. Jeffrey N. Weitzel, Elizabeth C. Chao, Bita Nehoray, et al. Somatic TP53 variants frequently confound germ-line testing results. Genetics in Medicine, 20(8):809–816, 8 2018. ISSN 15300366. doi: 10.1038/gim.2017.196.

4. Thomas P. Slavin, Bradford Coffee, Ryan Bernhisel, et al. Prevalence and characteristics of likely-somatic variants in cancer susceptibility genes among individuals who had hereditary pan-cancer panel testing. Cancer Genetics, 235–236:31–38, 6 2019. ISSN 22107770. doi: 10.1016/j.cancergen.2019.04.005.

5. Bradford Coffee, Hannah C. Cox, Ryan Bernhisel, et al. A substantial proportion of apparently heterozygous TP53 pathogenic variants detected with a next-generation sequencing hereditary pan-cancer panel are acquired somatically. Human Mutation, 41(1), 2020. ISSN 10981004. doi: 10.1002/humu.23910.

6. Alison N. Schwartz, Sophie R. Cahill, Samantha M. Stokes, et al. Evaluation of TP53 Variants Detected on Peripheral Blood or Saliva Testing: Discerning Germline From Somatic TP53 Variants. JCO Precision Oncology, (5):1677–1686, 11 2021. doi:10.1200/PO.21.00278.

7. Ozge Ceyhan-Birsoy, Pier Selenica, M. Herman Chui, et al. Paired Tumor-Normal Sequencing Provides Insights Into the TP53-Related Cancer Spectrum in Patients With Li-Fraumeni Syndrome. JNCI: Journal of the National Cancer Institute, 113(12):1751–1760, 11 2021. ISSN 0027-8874. doi: 10.1093/JNCI/DJAB117.

8. Paula Rofes, Carmen Castillo-Manzano, Mireia Menéndez, et al. TP53 germline testing and hereditary cancer: how somatic events and clinical criteria affect variant detection rate. Genome medicine, 17(1):3, 1 2025. ISSN 1756994X. doi: 10.1186/S13073-025-01429-5/FIGURES/4.

9. Hamish A J MacGregor, Duc Tran, Kelly L Bolton, et al. The age-related deceleration of clonal haematopoiesis in 420,000 healthy adults. bioRxiv, page 2023.12.21.572706, 3 2025. doi: 10.1101/2023.12.21.572706.

10. Giulio Genovese, Anna K. Kähler, Robert E. Handsaker, et al. Clonal Hematopoiesis and Blood-Cancer Risk Inferred from Blood DNA Sequence. New England Journal of Medicine, 371(26):2477–2487, 12 2014.

11. Siddhartha Jaiswal, Pierre Fontanillas, Jason Flannick, et al. Age-Related Clonal Hematopoiesis Associated with Adverse Outcomes. New England Journal of Medicine, 371(26), 12 2014. ISSN 0028-4793. doi: 10.1056/NEJMoa1408617.

12. Mingchao Xie, Charles Lu, Jiayin Wang, et al. Age-related mutations associated with clonal hematopoietic expansion and malignancies. Nature Medicine, 20(12):1472–1478, 12 2014. ISSN 1546170X. doi: 10.1038/NM.3733.

13. Thomas McKerrell, Naomi Park, Thaidy Moreno, et al. Leukemia-Associated Somatic Mutations Drive Distinct Patterns of Age-Related Clonal Hemopoiesis. Cell Reports, 10(8): 1239–1245, 3 2015.

14. Muxin Gu, Sruthi Cheloor Kovilakam, William G. Dunn, et al. Multiparameter prediction of myeloid neoplasia risk. Nature Genetics 2023 55:9, 55(9):1523–1530, 8 2023. ISSN 1546-1718. doi: 10.1038/s41588-023-01472-1.

15. Daniel A. Arber, Attilio Orazi, Robert P. Hasserjian, et al. International Consensus Classification of Myeloid Neoplasms and Acute Leukemias: integrating morphologic, clinical, and genomic data. Blood, 140(11):1200–1228, 9 2022. ISSN 0006-4971. doi: 10.1182/BLOOD.2022015850.

16. Moazzam Shahzad, Muhammad Kashif Amin, Naval G. Daver, et al. What have we learned about TP53-mutated acute myeloid leukemia? Blood Cancer Journal 2024 14:1, 14(1):1–9, 11 2024. ISSN 2044-5385. doi: 10.1038/s41408-024-01186-5.

17. David Malkin. Li-Fraumeni Syndrome. Genes & Cancer, 2(4):475, 4 2011. doi: 10.1177/1947601911413466.

18. A Chompret, L Brugières, M Ronsin, et al. P53 germline mutations in childhood cancers and cancer risk for carrier individuals. British Journal of Cancer 2000 82:12, 82(12):1932–1937, 5 2000. ISSN 1532-1827. doi: 10.1054/bjoc.2000.1167.

19. Ana Beatriz Sánchez-Heras, Teresa Ramon y Cajal, Marta Pineda, et al. SEOM clinical guideline on heritable TP53-related cancer syndrome (2022). Clinical and Translational Oncology, 25(9):2627–2633, 9 2023. ISSN 16993055. doi: 10.1007/S12094-023-03202-9/TABLES/2.

20. Maria Isabel Achatz and Gerard P. Zambetti. The Inherited p53 Mutation in the Brazilian Population. Cold Spring Harbor Perspectives in Medicine, 6(12):a026195, 2016. ISSN 21571422. doi: 10.1101/CSHPERSPECT.A026195.

21. Jacquelyn Powers, Emilia M. Pinto, Thibaut Barnoud, et al. A rare TP53 mutation predominant in ashkenazi jews confers risk of multiple cancers. Cancer Research, 80(17): 3732–3744, 9 2020. ISSN 15387445. doi: 10.1158/0008-5472.CAN-20-1390/654534/AM/A-RARE-TP53-MUTATION-PREDOMINANT-IN-ASHKENAZI-JEWS.

22. David C. Stieg, Kaitlyn Casey, Bhanu Chandra Karisetty, et al. The Ashkenazi-Centric G334R Variant of TP53 is Severely Impaired for Transactivation but Retains Tumor Suppressor Function in a Mouse Model. Molecular and cellular biology, 44(12), 2024. ISSN 1098-5549. doi: 10.1080/10985549.2024.2421885.

23. Claire Freycon, Laura Palma, Crystal Budd, et al. Germline p.R181H variant in TP53 in a family exemplifying the genotype-phenotype correlations in Li-Fraumeni syndrome. Familial Cancer, 23(4):665–669, 11 2024. ISSN 15737292. doi: 10.1007/S10689-024-00419-7/TABLES/1.

24. Iñigo Martincorena, Amit Roshan, Moritz Gerstung, et al. High burden and pervasive positive selection of somatic mutations in normal human skin. Science, 348(6237):880–886, 5 2015. ISSN 0036-8075. doi: 10.1126/SCIENCE.AAA6806.

25. Iñigo Martincorena, Joanna C. Fowler, Agnieszka Wabik, et al. Somatic mutant clones colonize the human esophagus with age. Science, 362(6417):911–917, 11 2018. ISSN 10959203. doi: 10.1126/SCIENCE.AAU3879.

26. Emilie Abby, Stefan C. Dentro, Michael W.J. Hall, et al. Notch1 mutations drive clonal expansion in normal esophageal epithelium but impair tumor growth. Nature Genetics 2023 55:2, 55(2):232–245, 1 2023. ISSN 1546-1718. doi: 10.1038/s41588-022-01280-z.

27. William Hill, Emilia L. Lim, Clare E. Weeden, et al. Lung adenocarcinoma promotion by air pollutants. Nature 2023 616:7955, 616(7955):159–167, 4 2023. ISSN 1476-4687. doi: 10.1038/s41586-023-05874-3.

28. Humam Kadara, Smruthy Sivakumar, Yasminka Jakubek, et al. Driver mutations in normal airway epithelium elucidate spatiotemporal resolution of lung cancer. American Journal of Respiratory and Critical Care Medicine, 200(6):742–750, 9 2019. ISSN 15354970. doi:10.1164/RCCM.201806-1178OC/SUPPL{\_}FILE/DISCLOSURES.PDF.

29. John S. Welch, Timothy J. Ley, Daniel C. Link, et al. The origin and evolution of mutations in acute myeloid leukemia. Cell, 150(2):264–278, 7 2012. ISSN 10974172. doi: 10.1016/J.CELL.2012.06.023/ATTACHMENT/E87ACF10-71D2-41B4-B49E-F241231EE6CA/MMC6.XLS.

30. Cristina Fortuno, Kelly McGoldrick, Tina Pesaran, et al. Suspected clonal hematopoiesis as a natural functional assay of TP53 germline variant pathogenicity. Genetics in Medicine, 24 (3):673–680, 3 2022. ISSN 1098-3600. doi: 10.1016/J.GIM.2021.10.018.

31. Cristina Fortuno, Tina Pesaran, Jill Dolinsky, et al. An updated quantitative model to classify missense variants in the TP53 gene: A novel multifactorial strategy. Human Mutation, 42 (10):1351–1361, 10 2021. ISSN 10981004. doi: 10.1002/HUMU.24264.

32. Clare Bycroft, Colin Freeman, Desislava Petkova, et al. The UK Biobank resource with deep phenotyping and genomic data. Nature 2018 562:7726, 562(7726):203–209, 10 2018. ISSN 1476-4687. doi: 10.1038/s41586-018-0579-z.

33. Juba Nait Saada, Georgios Kalantzis, Derek Shyr, et al. Identity-by-descent detection across 487,409 British samples reveals fine scale population structure and ultra-rare variant associations. Nature Communications 2020 11:1, 11(1):1–15, 11 2020. ISSN 2041-1723. doi: 10.1038/s41467-020-19588-x.

34. Kelly L. Bolton, Ryan N. Ptashkin, Teng Gao, et al. Cancer therapy shapes the fitness landscape of clonal hematopoiesis. Nature Genetics, 52(11):1219–1226, 11 2020. ISSN 15461718. doi: 10.1038/s41588-020-00710-0.

35. Alexander Sun, Zhang 1, Meis Omran, et al. TP53 p.R181H is enriched in the Swedish cohort (SWEP53) and associated with a distinct breast and prostate phenotype. Scientific Reports 2025 15:1, 15(1):35033–, 10 2025. ISSN 2045-2322. doi: 10.1038/s41598-025-22407-2.

36. Andrew O. Giacomelli, Xiaoping Yang, Robert E. Lintner, et al. Mutational processes shape the landscape of TP53 mutations in human cancer. Nature Genetics 2018 50:10, 50(10): 1381–1387, 9 2018. ISSN 1546-1718. doi: 10.1038/s41588-018-0204-y.

37. Gaëlle Bougeard, Mariette Renaux-Petel, Jean Michel Flaman, et al. Revisiting Li-Fraumeni syndrome from TP53 mutation carriers. Journal of Clinical Oncology, 33 (21):2345–2352, 7 2015. ISSN 15277755. doi: 10.1200/JCO.2014.59.5728/ASSET/A7B0C07C-564D-4EEA-850E-BAD9613CA386/ASSETS/GRAPHIC/ZLJ9991052740006. JPEG.

38. The TP53 Database (R21, Jan 2025).

39. Kelvin César de Andrade, Elaine E. Lee, Elise M. Tookmanian, et al. The TP53 Database: transition from the International Agency for Research on Cancer to the US National Cancer Institute. Cell Death and Differentiation, 29(5):1071–1073, 5 2022. ISSN 14765403. doi: 10.1038/S41418-022-00976-3.

40. Caroline J. Watson, A. L. Papula, Gladys Y. P. Poon, et al. The evolutionary dynamics and fitness landscape of clonal hematopoiesis. Science, 367(6485):1449–1454, 3 2020. ISSN 0036-8075. doi: 10.1126/SCIENCE.AAY9333.

41. Naval G. Daver, Abhishek Maiti, Tapan M. Kadia, et al. TP53-Mutated Myelodysplastic Syndrome and Acute Myeloid Leukemia: Biology, Current Therapy, and Future Directions. Cancer Discovery, 12(11):2516–2529,11 2022. ISSN 21598290. doi: 10.1158/2159-8290.CD-22-0332/709691/P/TP53-MUTATED-MYELODYSPLASTIC-SYNDROME-AND-ACUTE.

42. Caroline J. Watson, Gladys Y. P. Poon, Hamish A.J. MacGregor, et al. Evolutionary dynamics in the decades preceding acute myeloid leukaemia. bioRxiv, page 2024.07.05.602251, 7 2024. doi: 10.1101/2024.07.05.602251.

43. Siddhartha P. Kar, Pedro M. Quiros, Muxin Gu, et al. Genome-wide analyses of 200,453 individuals yield new insights into the causes and consequences of clonal hematopoiesis. Nature Genetics 2022 54:8, 54(8):1155–1166, 7 2022. ISSN 1546-1718. doi: 10.1038/s41588-022-01121-z.

44. Zhihui Xi, Huolun Feng, Kunling Chen, et al. Clonal hematopoiesis of indeterminate potential is a risk factor of gastric cancer: A Prospective Cohort in UK Biobank study. Translational Oncology, 52:102242, 2 2025. ISSN 1936-5233. doi: 10.1016/J.TRANON.2024.102242.

45. Yongfeng Liu, Zhihui Xi, Jianlong Zhou, et al. Clonal Hematopoiesis of Indeterminate Potential as a Predictor of Colorectal Cancer Risk: Insights from the UK Biobank Cohort. Cancer epidemiology, biomarkers & prevention : a publication of the American Association for Cancer Research, cosponsored by the American Society of Preventive Oncology, 34 (3):405–411, 3 2025. ISSN 15387755. doi: 10.1158/1055-9965.EPI-24-1342/751072/AM/CLONAL-HEMATOPOIESIS-OF-INDETERMINANT-POTENTIAL-AS.

46. Pinkal Desai, Ying Zhou, Justin Grenet, et al. Association of clonal hematopoiesis and mosaic chromosomal alterations with solid malignancy incidence and mortality. Cancer, 130(22):3879–3887, 11 2024. ISSN 1097-0142. doi: 10.1002/CNCR.35455.

47. Cancer incidence for all cancers combined | Cancer Research UK.

48. Kristian Cibulskis, Michael S. Lawrence, Scott L. Carter, et al. Sensitive detection of somatic point mutations in impure and heterogeneous cancer samples. Nature Biotechnology 2013 31:3, 31(3):213–219, 2 2013. ISSN 1546-1696. doi: 10.1038/nbt.2514.

49. Duc Tran, J. Scott Beeler, Jie Liu, et al. Plasma Proteomic Signature Predicts Myeloid Neoplasm Risk. Clinical Cancer Research, 30(15):3220–3228, 8 2024. ISSN 15573265. doi: 10.1158/1078-0432.CCR-23-3468/735081/AM/PLASMA-PROTEOMIC-SIGNATURE-PREDICTS-MYELOID.

50. Vikas Bansal and Ondrej Libiger. Fast individual ancestry inference from DNA sequence data leveraging allele frequencies for multiple populations. BMC Bioinformatics, 16(1):1–11, 1 2015. ISSN 14712105. doi: 10.1186/S12859-014-0418-7/TABLES/2.

51. Lorenzo Ficorella, Xin Yang, Nasim Mavaddat, et al. Adapting the BOADICEA breast and ovarian cancer risk models for the ethnically diverse UK population. British Journal of Cancer 2025 133:6, 133(6):844–855, 7 2025. ISSN 1532-1827. doi: 10.1038/s41416-025-03117-y.

52. Mao Jan Lin, Sheila Iyer, Nae Chyun Chen, and Ben Langmead. Measuring, visualizing, and diagnosing reference bias with biastools. Genome Biology, 25(1):1–28, 12 2024. ISSN 1474760X. doi: 10.1186/S13059-024-03240-8/FIGURES/9.

53. Clare Bycroft, Colin Freeman, Desislava Petkova, et al. Genome-wide genetic data on ∼500,000 UK Biobank participants. bioRxiv, page 166298, 7 2017. doi: 10.1101/166298.

54. Jared O’Connell, Kevin Sharp, Nick Shrine, et al. Haplotype estimation for biobank-scale data sets. Nature Genetics 2016 48:7, 48(7):817–820, 6 2016. ISSN 1546-1718. doi: 10.1038/ng.3583.

55. Adam Auton, Gonçalo R. Abecasis, David M. Altshuler, et al. A global reference for human genetic variation. Nature 2015 526:7571, 526(7571):68–74, 9 2015. ISSN 1476-4687. doi: 10.1038/nature15393.

56. Bjarni V. Halldorsson, Gunnar Palsson, Olafur A. Stefansson, et al. Human genetics: Characterizing mutagenic effects of recombination through a sequence-level genetic map. Science, 363(6425), 1 2019. ISSN 10959203. doi: 10.1126/SCIENCE.AAU1043/SUPPL{\_}FILE/AAU1043{\_}DATAS7.TSV.

57. Po Ru Loh, Giulio Genovese, Robert E. Handsaker, et al. Insights into clonal haematopoiesis from 8,342 mosaic chromosomal alterations. Nature, 559(7714):350–355, 7 2018. ISSN 14764687. doi: 10.1038/S41586-018-0321-X.

58. Jessica L. Mester, Sarah A. Jackson, Kristen Postula, et al. Apparently Heterozygous TP53 Pathogenic Variants May Be Blood Limited in Patients Undergoing Hereditary Cancer Panel Testing. Journal of Molecular Diagnostics, 22(3):396–404, 3 2020. ISSN 19437811. doi: 10.1016/j.jmoldx.2019.12.003.

59. Mariette Renaux-Petel, Françoise Charbonnier, Jean Christophe Théry, et al. Contribution of de novo and mosaic TP53 mutations to Li-Fraumeni syndrome. Journal of Medical Genetics, 55(3):173–180, 3 2018. ISSN 0022-2593. doi: 10.1136/JMEDGENET-2017-104976.

60. Skipper Seabold and Josef Perktold. Statsmodels: Econometric and Statistical Modeling with Python. Proceedings of the 9th Python in Science Conference, pages 92–96, 1 2010. doi: 10.25080/MAJORA-92BF1922-011.

61. Cameron Davidson-Pilon. lifelines, survival analysis in Python. doi: 10.5281/ZENODO.14007206.

